# Distinct workforce activation states among licensed US nurses outside nursing employment

**DOI:** 10.64898/2026.09.19.26363485

**Authors:** Meina Huang, Zheyi Chen

## Abstract

**Background:** Licensed registered nurses (RNs) who are not employed in nursing are often counted as reserve capacity. A license, however, does not indicate readiness to enter the workforce. We examined whether this population contains distinct workforce activation states and how reported reasons for not working in nursing correspond to those states.

**Methods:** We analyzed the 2022 National Sample Survey of Registered Nurses, a nationally representative cross-sectional survey of licensed US nurses. The study population comprised RNs and RN–advanced practice registered nurses who were actively licensed but not working for pay in nursing on 31 December 2021. The outcome distinguished five states reported at survey response: no future intention, actively looking, plans future entry or return, undecided, and already entered or returned. Ten prespecified reasons with adequate cell sizes were examined in separate survey-weighted multinomial models. Variance estimation used the main survey weight and 80 successive-difference replicate weights. Models adjusted for age, sex, race and ethnicity, nursing education and license type; the family-caregiving model also adjusted for marital status and dependents.

**Results:** The model-eligible sample included 7,744 respondents, representing 869,236 licensed RNs. An estimated 44.4% had no future intention to work in nursing, 25.6% were undecided, 14.6% had entered or returned, 10.7% planned future entry or return, and 4.7% were actively looking. Retirement was associated with a 27.0-percentage-point higher adjusted probability of no future intention (95% CI 21.1 to 32.9). Family caregiving was associated with a 14.6-point higher probability of planned entry or return (95% CI 10.9 to 18.3) and a 13.3-point lower probability of no future intention (95% CI −18.0 to −8.5). Burnout, stressful work, inadequate staffing, scheduling problems and poor management were each concentrated in the undecided state, with adjusted probability differences from 6.1 to 14.6 points. These patterns remained in analyses restricted by retirement, prior nursing work, license type and official imputation status.

**Conclusions:** The licensed nonworking RN population is a stratified labor reserve. Family caregiving identifies a planned reserve, workplace barriers identify a conditional reserve, and retirement identifies a largely non-mobilizable group. Workforce planning should measure activation state before treating licensed capacity as available supply.

## Introduction

Health-workforce policy depends on the conversion of trained and licensed capacity into effective labor supply. The World Health Organization, International Labour Organization and Organisation for Economic Co-operation and Development distinguish gaining employment, maintaining employment, improving working conditions, training and retention as separate components of health and care workforce policy [1]. This distinction has become more important since the COVID-19 pandemic. US nurse employment fell sharply during the early pandemic and later rebounded, yet aggregate growth and full-time-equivalent projections do not resolve shortages by setting, geography or working conditions [2,3]. Post-pandemic policy therefore requires attention to retention, distribution and working conditions alongside educational expansion [4].

One possible source of additional supply is the population of actively licensed RNs who are not employed in nursing. Official 2022 National Sample Survey of Registered Nurses (NSSRN) reports describe widespread burnout and dissatisfaction, pandemic-related exits and heterogeneous intentions among nurses who left during the pandemic [5,6]. Analysis of the 2018 NSSRN estimated substantial reserve capacity among licensed RNs not working in nursing, while noting that the data did not establish their willingness, ability, preparation needs or required accommodations [7]. A professional license is therefore a measure of potential capacity, not immediate availability.

Earlier studies also show that exit from nursing is heterogeneous. National NSSRN analyses distinguish career change from labor-force separation and identify family structure, education and age as different correlates of those pathways [8,9]. More recent survey evidence indicates that retirement, burnout, insufficient staffing and family obligations often coexist when nurses end health-care employment [10]. Studies of inactive nurses have identified career commitment, schedules, parenting demands, workload, safe conditions and re-entry preparation as potential influences on return [11–13]. Pandemic experience further showed that a licensed inactive pool cannot be assumed to be immediately deployable when previous work experiences and perceived risks remain unresolved [14]. A recent synthesis frames nursing careers as movement through entry, retention, exit and return, with push and pull factors that change across career stages [15].

The stage of workforce activation matters because intention, active search and observed employment are not interchangeable. Classic turnover theory places intermediate cognitions and search behavior between job dissatisfaction and actual movement [16], and later work recognizes multiple withdrawal states, shocks and forms of embeddedness rather than a single linear pathway [17,18]. For licensed RNs outside nursing employment, being undecided, planning a later return, actively looking and already entering or returning represent different distances from effective supply. Collapsing these states into a binary willingness-to-return measure obscures the type of barrier and the policy response it may require.

Workplace conditions are plausible markers of these states. Inadequate staffing is associated with nurse burnout, dissatisfaction and patient outcomes [19], and chronic understaffing compounded adverse safety and workforce assessments during the pandemic [20]. Staffing policy can change workloads and patient outcomes [21], while long shifts and overtime are associated with burnout, dissatisfaction and turnover [22–24]. Burnout itself is multidimensional and linked to workload, the practice environment and employment outcomes [25–27]. Leadership, supervisory support, work-family conflict and team cohesion are also prominent correlates of turnover and retention [28–31]. Workplace violence and harassment add a distinct occupational hazard [32–34]. By contrast, family caregiving and retirement may signal time constraints or life-course transitions rather than rejection of nursing work. Work-family conflict is associated with turnover intention [35], and nurses providing care to children or older relatives identify flexibility as an important workplace condition [36–38]. The timing of retirement likewise reflects health, work ability, income, flexibility and organizational recognition [39,40].

We used the 2022 NSSRN to characterize workforce activation among actively licensed RNs outside nursing employment. The study addressed three questions: how this population is distributed across five activation states; how personal, family, health and workplace reasons correspond to those states; and whether the associations persist across alternative definitions of the potentially mobilizable population. We expected retirement to concentrate in the no-future-intention state, family caregiving to concentrate in planned future entry or return, and workplace barriers to concentrate among nurses whose entry or return remained undecided.

## Methods

### Study design and data source

This cross-sectional study used the state-based public-use file of the 2022 NSSRN [41]. The National Center for Health Workforce Analysis within the Health Resources and Services Administration (HRSA) sponsored the survey, and the US Census Bureau administered it [42]. Data collection occurred from 15 December 2022 through 13 April 2023. The probability sample covered the 50 states and the District of Columbia and represented US-resident nurses who held an active RN license on 31 December 2021.

Within each state, sampling distinguished nurse practitioners from other RNs and used differential sampling rates. The survey collected information on demographic characteristics, education, employment, earnings, working conditions and future employment plans. The analysis followed the Strengthening the Reporting of Observational Studies in Epidemiology (STROBE) guidance for cross-sectional studies [43,44].

### Study population

The public-use file contained 49,114 respondents. We included respondents whose license status identified them as an RN or an RN and advanced practice registered nurse (APRN) and who reported that they were not working for pay in nursing on 31 December 2021. This definition produced a target sample of 7,753 respondents. Nine respondents were excluded from adjusted analyses because highest nursing education was missing, leaving 7,744 model-eligible respondents. The study population included both nurses with previous nursing employment and 774 respondents who had never worked in nursing. We therefore use *entry or return* for the full population and reserve *return* for those with previous nursing employment.

### Workforce activation states

The outcome was derived from the survey item asking nonworking nurses about their current or future nursing employment status. We defined five mutually exclusive states: (1) no future intention to work for pay in nursing; (2) actively looking for nursing work; (3) plans future entry or return; (4) undecided; and (5) already entered or returned after 31 December 2021. The first category was the reference outcome in multinomial models. The five-state formulation preserves the distinction between latent interest, active job search and realized employment.

### Reasons for not working in nursing

The survey allowed respondents to select any of 23 reasons for not working in nursing. We retained all reasons for descriptive analysis. They covered retirement; family and health constraints; physical demands; burnout and stress; staffing, scheduling, pay, management, collaboration, advancement and safety; liability and professional-practice concerns; career change and education; outdated skills and job matching; and other reasons. Table 2 reports every item separately.

**Table 1.** Characteristics of the model-eligible population by workforce activation state. Values are unweighted n (survey-weighted percentage [95% CI]) unless otherwise indicated. Weighted estimates use the NSSRN main weight and all 80 successive-difference replicate weights.

| Characteristic | Level | Overall | No future intention | Actively looking | Planned entry/return | Undecided | Entered/returned |
| --- | --- | --- | --- | --- | --- | --- | --- |
| Sample size |  | 7,744; weighted N=869,236 | 3,696; weighted N=386,029 | 292; weighted N=40,553 | 697; weighted N=93,060 | 2,086; weighted N=222,678 | 973; weighted N=126,917 |
| Age, years | <50 | 1,771 (32.0% [30.3-33.7]) | 271 (12.2% [10.0-14.5]) | 118 (50.5% [40.9-60.1]) | 379 (63.0% [57.7-68.3]) | 411 (25.7% [22.4-29.0]) | 592 (74.9% [70.2-79.5]) |
| Age, years | 50-64 | 2,009 (26.8% [25.2-28.4]) | 861 (26.4% [23.9-29.0]) | 93 (36.8% [27.6-46.0]) | 180 (22.5% [17.4-27.6]) | 632 (31.6% [28.1-35.2]) | 243 (19.5% [15.2-23.7]) |
| Age, years | 65+ | 3,964 (41.2% [39.5-42.8]) | 2,564 (61.4% [58.8-64.0]) | 81 (12.6% [7.8-17.5]) | 138 (14.5% [10.7-18.3]) | 1,043 (42.6% [39.2-46.1]) | 138 (5.7% [3.8-7.5]) |
| Sex | Male | 528 (8.8% [7.7-9.8]) | 223 (7.5% [6.1-8.9]) | 23 (15.2% [5.7-24.7]) | 62 (13.4% [8.5-18.2]) | 125 (6.2% [4.7-7.7]) | 95 (11.8% [8.3-15.3]) |
| Sex | Female | 7,216 (91.2% [90.2-92.3]) | 3,473 (92.5% [91.1-93.9]) | 269 (84.8% [75.3-94.3]) | 635 (86.6% [81.8-91.5]) | 1,961 (93.8% [92.3-95.3]) | 878 (88.2% [84.7-91.7]) |
| Race/ethnicity | White, non-Hispanic | 6,709 (74.8% [72.9-76.7]) | 3,355 (80.3% [77.7-83.0]) | 218 (54.1% [41.8-66.5]) | 554 (66.6% [60.8-72.4]) | 1,812 (79.1% [75.9-82.3]) | 770 (63.3% [56.7-69.8]) |
| Race/ethnicity | Black, non-Hispanic | 304 (8.3% [7.1-9.6]) | 112 (6.8% [5.1-8.6]) | 25 (11.9% [4.8-19.0]) | 32 (8.3% [3.9-12.8]) | 83 (9.4% [7.0-11.9]) | 52 (9.7% [5.3-14.1]) |
| Race/ethnicity | Hispanic | 212 (5.5% [4.3-6.6]) | 57 (2.9% [1.6-4.2]) | 18 (20.1% [8.7-31.6]) | 27 (5.9% [2.2-9.5]) | 56 (3.2% [1.8-4.6]) | 54 (12.0% [7.9-16.2]) |
| Race/ethnicity | Other | 519 (11.4% [10.0-12.9]) | 172 (9.9% [7.7-12.1]) | 31 (13.8% [5.7-21.9]) | 84 (19.2% [13.4-25.1]) | 135 (8.3% [5.7-10.9]) | 97 (15.0% [11.0-18.9]) |
| Highest nursing education | BSN | 2,642 (46.7% [44.7-48.8]) | 1,130 (41.7% [38.6-44.7]) | 101 (56.6% [46.3-66.8]) | 305 (58.8% [52.6-65.0]) | 697 (41.4% [37.9-44.9]) | 409 (59.6% [54.4-64.8]) |
| Highest nursing education | Diploma/ADN | 2,500 (39.3% [37.6-41.1]) | 1,312 (42.4% [39.4-45.4]) | 79 (33.6% [23.9-43.3]) | 170 (30.8% [24.8-36.9]) | 712 (45.0% [41.4-48.7]) | 227 (28.0% [23.1-33.0]) |
| Highest nursing education | Graduate | 2,602 (13.9% [12.8-15.0]) | 1,254 (15.9% [13.8-18.0]) | 112 (9.9% [6.0-13.7]) | 222 (10.4% [6.2-14.5]) | 677 (13.6% [11.4-15.8]) | 337 (12.4% [9.3-15.5]) |
| Marital status | Married/domestic partnership | 5,623 (72.3% [70.5-74.1]) | 2,710 (74.1% [71.6-76.7]) | 205 (65.0% [53.3-76.6]) | 543 (80.2% [75.2-85.2]) | 1,500 (71.4% [68.5-74.4]) | 665 (64.8% [60.0-69.5]) |
| Marital status | Widowed/divorced/separated | 1,548 (18.2% [16.8-19.7]) | 794 (19.7% [17.7-21.7]) | 58 (19.4% [9.5-29.3]) | 87 (9.7% [5.4-14.0]) | 461 (21.2% [18.4-23.9]) | 148 (14.7% [11.0-18.5]) |
| Marital status | Never married | 573 (9.5% [8.3-10.6]) | 192 (6.2% [4.6-7.7]) | 29 (15.6% [6.9-24.4]) | 67 (10.1% [6.2-14.0]) | 125 (7.4% [5.3-9.5]) | 160 (20.5% [16.7-24.3]) |
| Dependents | No dependents | 4,737 (55.5% [53.4-57.6]) | 2,721 (69.3% [66.3-72.3]) | 143 (40.7% [29.8-51.7]) | 243 (32.5% [26.6-38.3]) | 1,185 (54.3% [50.5-58.1]) | 445 (37.5% [32.2-42.8]) |
| Dependents | Any dependents | 3,007 (44.5% [42.4-46.6]) | 975 (30.7% [27.7-33.7]) | 149 (59.3% [48.3-70.2]) | 454 (67.5% [61.7-73.4]) | 901 (45.7% [41.9-49.5]) | 528 (62.5% [57.2-67.8]) |
| Nursing licensure type | RN only | 5,336 (94.0% [93.4-94.7]) | 2,599 (93.6% [92.6-94.6]) | 182 (94.8% [93.1-96.5]) | 480 (96.2% [95.4-96.9]) | 1,424 (93.5% [92.4-94.5]) | 651 (94.6% [93.5-95.6]) |
| Nursing licensure type | RN and APRN | 2,408 (6.0% [5.3-6.6]) | 1,097 (6.4% [5.4-7.4]) | 110 (5.2% [3.5-6.9]) | 217 (3.8% [3.1-4.6]) | 662 (6.5% [5.5-7.6]) | 322 (5.4% [4.4-6.5]) |
| S. Census region of residence | Northeast | 1,567 (22.2% [20.7-23.7]) | 797 (24.2% [21.7-26.8]) | 63 (23.1% [14.4-31.8]) | 115 (14.7% [10.8-18.6]) | 430 (22.6% [19.7-25.5]) | 162 (20.9% [16.5-25.2]) |
| S. Census region of residence | Midwest | 1,915 (25.1% [23.7-26.5]) | 987 (27.7% [25.3-30.0]) | 51 (11.6% [6.3-16.9]) | 138 (18.4% [14.7-22.1]) | 503 (26.1% [23.5-28.8]) | 236 (24.7% [20.4-29.0]) |
| S. Census region of residence | South | 2,380 (35.1% [33.3-36.8]) | 1,052 (32.5% [29.9-35.1]) | 95 (28.3% [20.2-36.4]) | 240 (43.7% [36.8-50.5]) | 669 (37.5% [34.1-40.9]) | 324 (34.4% [29.3-39.5]) |
| S. Census region of residence | West | 1,882 (17.6% [16.4-18.8]) | 860 (15.6% [13.7-17.5]) | 83 (37.0% [25.1-48.9]) | 204 (23.2% [17.7-28.7]) | 484 (13.8% [11.3-16.2]) | 251 (20.0% [15.0-25.1]) |

**Table 2.**
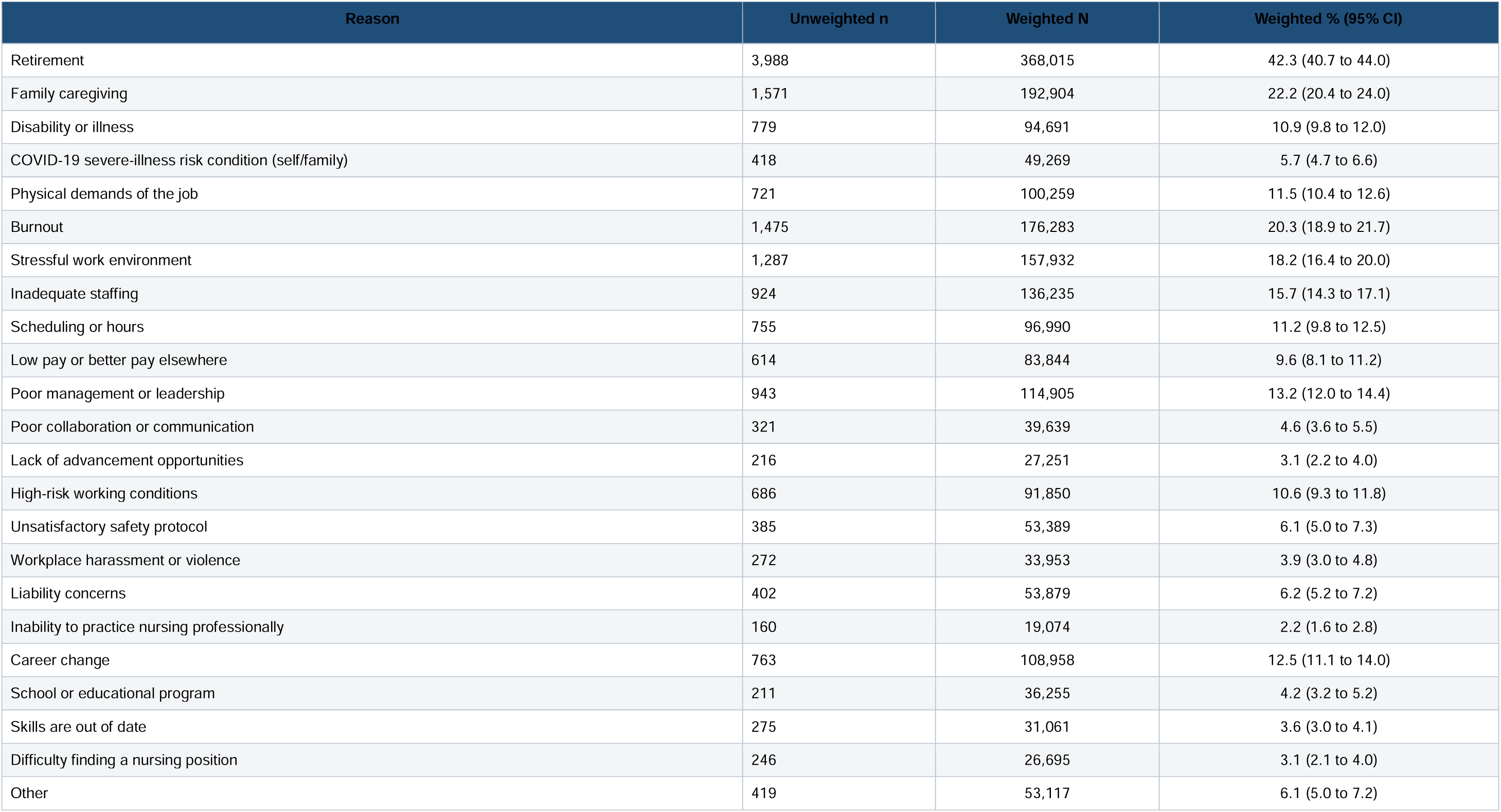
Reported reasons for not working in nursing. All 23 official survey reasons are shown. Percentages and confidence intervals are survey weighted; reasons were not mutually exclusive.

Inferential analyses were prespecified for reasons with at least 30 respondents in every reason-by-state cell. Ten reason domains met this threshold: retirement, family caregiving, health or physical limitations, burnout, stressful work environment, inadequate staffing, scheduling or hours, low pay, poor management or leadership, and safety or violence. Health combined disability or illness with physical demands. Safety combined high-risk working conditions, unsatisfactory safety protocol and workplace harassment or violence. Reasons were not treated as mutually exclusive.

### Covariates

Covariates were chosen before modeling to describe demographic and professional differences likely to structure labor-force participation. All models adjusted for age, sex, race and ethnicity, highest nursing education and RN–APRN license status. Age was modeled with a natural spline with three degrees of freedom to accommodate its nonlinear association with retirement and employment. Race and ethnicity were grouped as Hispanic, non-Hispanic White, non-Hispanic Black and other race or ethnicity. Highest nursing education was grouped as diploma or associate degree, bachelor of science in nursing, and graduate nursing degree. The family-caregiving model additionally adjusted for marital status and the presence of dependents because these variables directly structure caregiving opportunities and constraints. Adjustment-variable coefficients were not interpreted as independent effects [45].

### Statistical analysis

National estimates used the final person weight supplied with the NSSRN public-use file. Variances and 95% confidence intervals used all 80 successive-difference replication (SDR) weight columns and the mean-squared-error formulation. For an estimate θ, the replicate variance was (4/80) times the sum of squared differences between each replicate estimate and the full-sample estimate. We created the replicate-weight design before defining the study population as a survey domain. Design-based inference was implemented with the *survey* package, which supports replicate-weight analysis of complex survey samples [46].

We first estimated weighted counts and proportions for activation states, respondent characteristics and all 23 reasons. Pairwise weighted phi correlations described co-occurrence among the ten modeled reasons. Because several workplace reasons were strongly correlated and respondents could select multiple reasons, we fitted a separate multinomial model for each reason instead of entering all reasons into one model. This approach estimates each reason’s adjusted population profile without assigning mutually adjusted interpretations to overlapping self-reported experiences.

Multinomial models were fitted to the main weight and refitted for every replicate weight. From each model, we reported relative risk ratios and reason-level four-degree-of-freedom joint tests across the four nonreference outcome logits. Joint Wald statistics were converted to finite-design F tests with 79 denominator degrees of freedom. To improve interpretability, we standardized predictions over the observed covariate distribution after setting the modeled reason to present and absent for every respondent. The principal estimands were adjusted marginal probabilities and their percentage-point differences. Benjamini–Hochberg correction controlled the false-discovery rate across the ten reason-level joint tests and, separately, across the 50 state-specific marginal contrasts. All tests were two-sided.

Five sensitivity analyses assessed stability. They excluded respondents who had already entered or returned; excluded those selecting retirement; excluded records with an officially imputed outcome or reason block; restricted the sample to non-APRN RNs; and restricted the sample to respondents with previous nursing work or observed entry or return. The main analysis retained official imputed survey values and used complete cases for model covariates. Analyses were conducted in R version 4.5.3 using *survey* version 4.5, *nnet* version 7.3-20 and *haven* version 2.5.5.

### Ethics and public involvement

In accordance with the authors’ institutional policy, this secondary analysis of a publicly available de-identified data file did not require local ethics committee review. The investigators had no interaction with participants, no access to direct identifiers and made no attempt to identify respondents. Participation in the original NSSRN was voluntary. The US Census Bureau protected responses under Title 13, United States Code, section 9; the federal information collection was approved under OMB Control No. 0607-1002. Additional informed consent was not required for this secondary analysis. No patients or members of the public were involved in setting the research question, conducting the analysis or preparing the manuscript.

## Results

### Population and activation states

The 7,744 model-eligible respondents represented 869,236 actively licensed RNs outside nursing employment. An estimated 386,029 (44.4%) had no future intention to work for pay in nursing, 222,678 (25.6%) were undecided, 126,917 (14.6%) had already entered or returned, 93,060 (10.7%) planned future entry or return, and 40,553 (4.7%) were actively looking (Figure 1a; Table 1).

**Figure 1.**
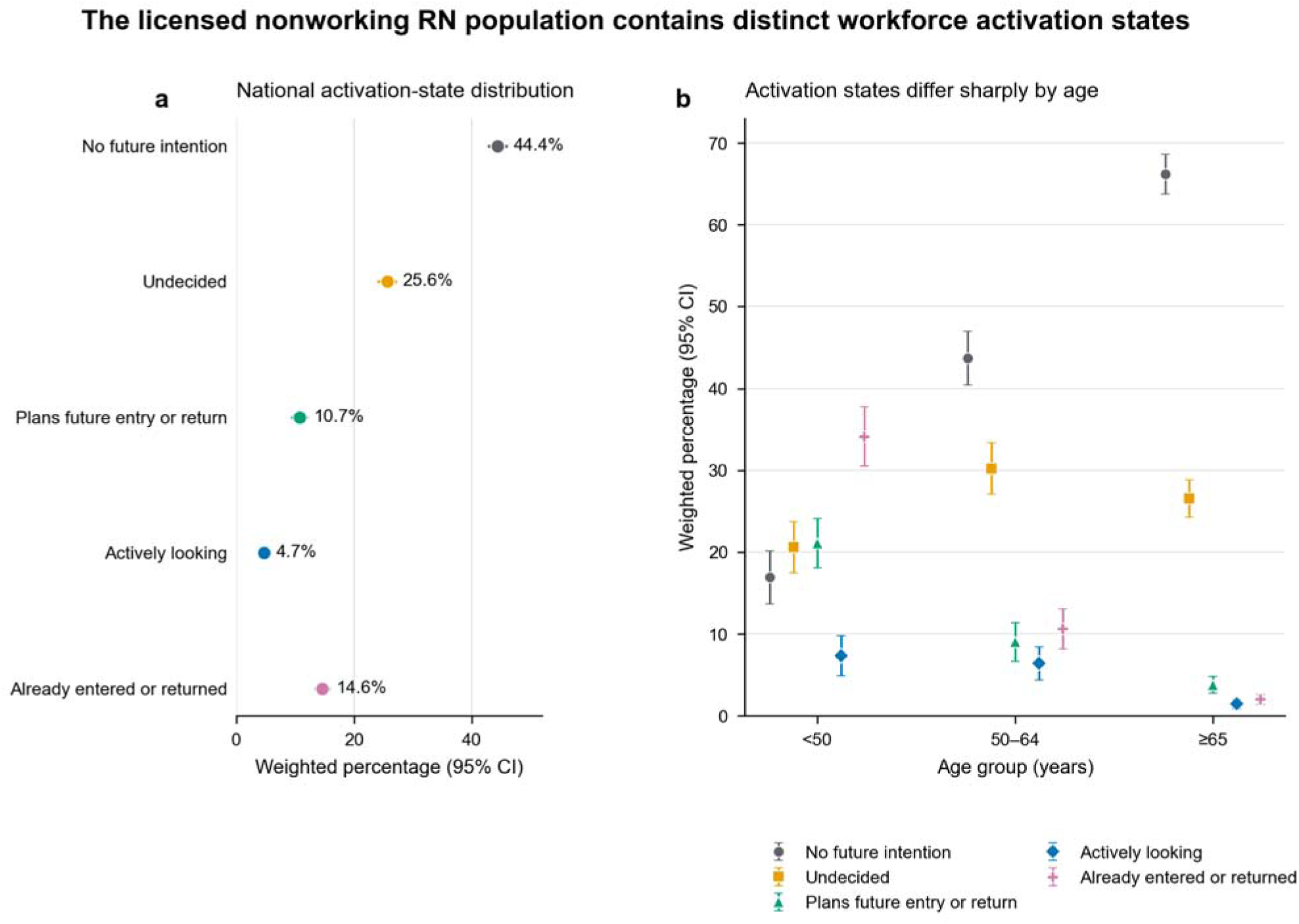
The licensed nonworking RN population contains distinct workforce activation states. **a**, Nationally weighted prevalence of five mutually exclusive activation states among actively licensed US RNs who were not working for pay in nursing on 31 December 2021. Points show percentages and horizontal lines show 95% confidence intervals. **b**, Weighted prevalence of the same states by age group. Estimates use the full target sample (n = 7,753) and the NSSRN main weight with successive-difference replicate weights. Entry includes respondents who had never worked in nursing; return refers to respondents with prior nursing employment.

Age sharply separated these states (Figure 1b). Among respondents younger than 50 years, 34.1% had already entered or returned and 21.0% planned future entry or return; 16.9% reported no future intention. Among those aged 65 years or older, 66.2% reported no future intention, 26.6% were undecided and 2.0% had already entered or returned. In the formal model sample, respondents younger than 50 years accounted for 74.9% of the already-entered-or-returned state and 63.0% of the planned-entry-or-return state, whereas respondents aged 65 years or older accounted for 61.4% of the no-future-intention state (Table 1).

The population was 91.2% female, 74.8% non-Hispanic White, and 94.0% licensed as an RN without an APRN license. Dependents were present for 44.5% overall but for 67.5% of the planned-entry-or-return state and 62.5% of the already-entered-or-returned state. These differences supported the planned adjustment for age, demographic characteristics, education and license type, with household variables added to the family-caregiving model.

### Reasons for not working in nursing

Retirement was the most frequently reported reason (42.3%, 95% CI 40.7% to 44.0%), followed by family caregiving (22.2%, 95% CI 20.4% to 24.0%), burnout (20.3%, 95% CI 18.9% to 21.7%), a stressful work environment (18.2%, 95% CI 16.4% to 20.0%), inadequate staffing (15.7%, 95% CI 14.3% to 17.1%), poor management or leadership (13.2%, 95% CI 12.0% to 14.4%), career change (12.5%, 95% CI 11.1% to 14.0%), physical demands (11.5%, 95% CI 10.4% to 12.6%), scheduling or hours (11.2%, 95% CI 9.8% to 12.5%) and high-risk working conditions (10.6%, 95% CI 9.3% to 11.8%) (Table 2). An estimated 59.1% reported one reason, 12.3% reported two and 28.7% reported three or more.

Workplace reasons clustered (Figure 3a). The strongest weighted phi correlations were poor management with inadequate staffing (0.59), burnout with stressful work (0.58), inadequate staffing with stressful work (0.57), and poor management with stressful work (0.55). Retirement was inversely correlated with family caregiving (−0.29) and with each modeled workplace reason (−0.15 to −0.22). This structure supported reason-specific rather than mutually adjusted models.

**Figure 2.**
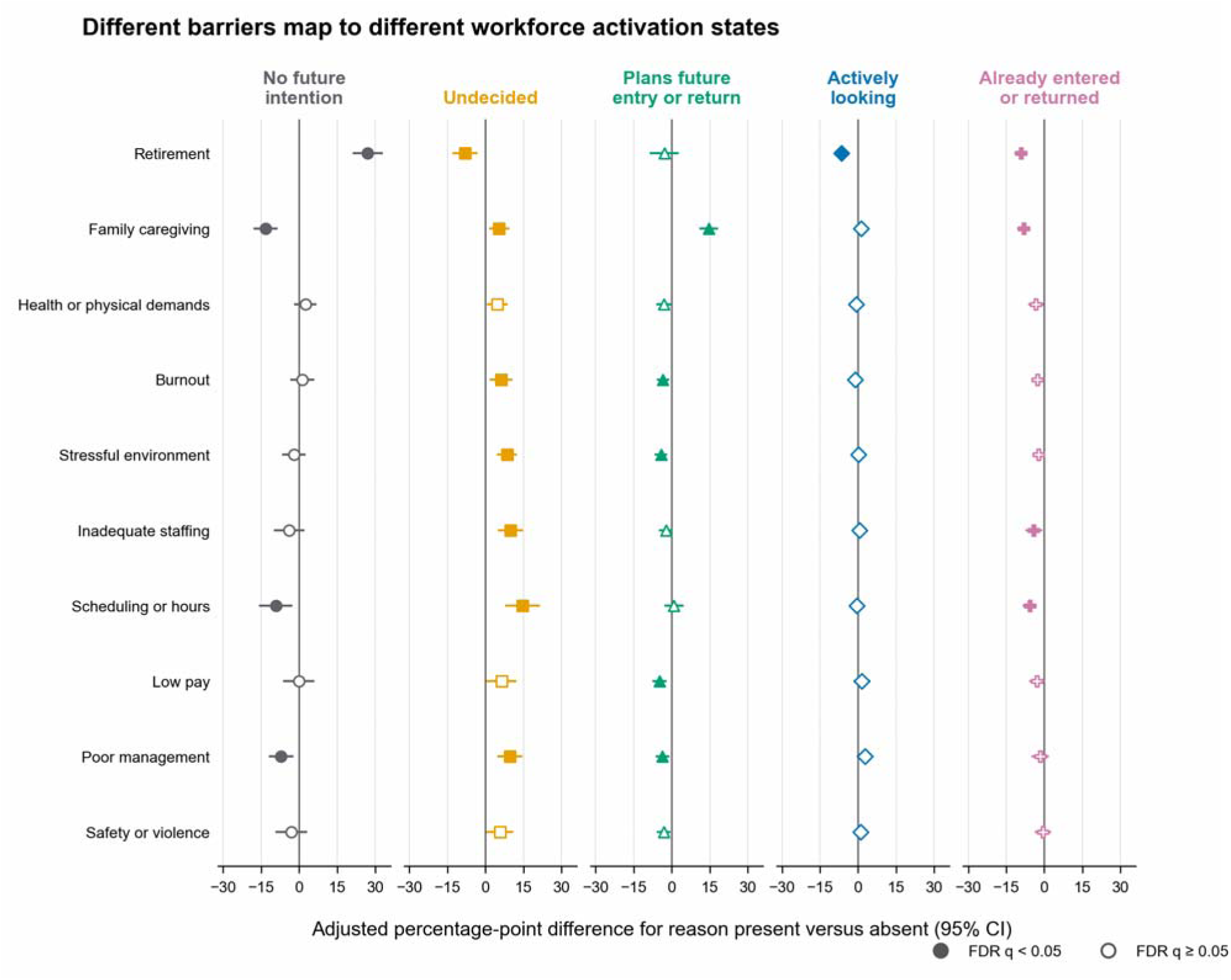
Different barriers map to different workforce activation states. Survey-weighted adjusted percentage-point differences in each activation state when a reported reason for not working in nursing was set present versus absent. Separate multinomial models were fitted for each reason among 7,744 model-eligible respondents. All models adjusted for age using a natural spline with three degrees of freedom, sex, race and ethnicity, highest nursing education and RN–APRN status; the family-caregiving model additionally adjusted for marital status and dependents. Lines show 95% confidence intervals. Filled symbols indicate contrast-level Benjamini–Hochberg false-discovery-rate q < 0.05 across the 50 marginal contrasts. Estimates describe associations and are not causal effects.

**Figure 3.**
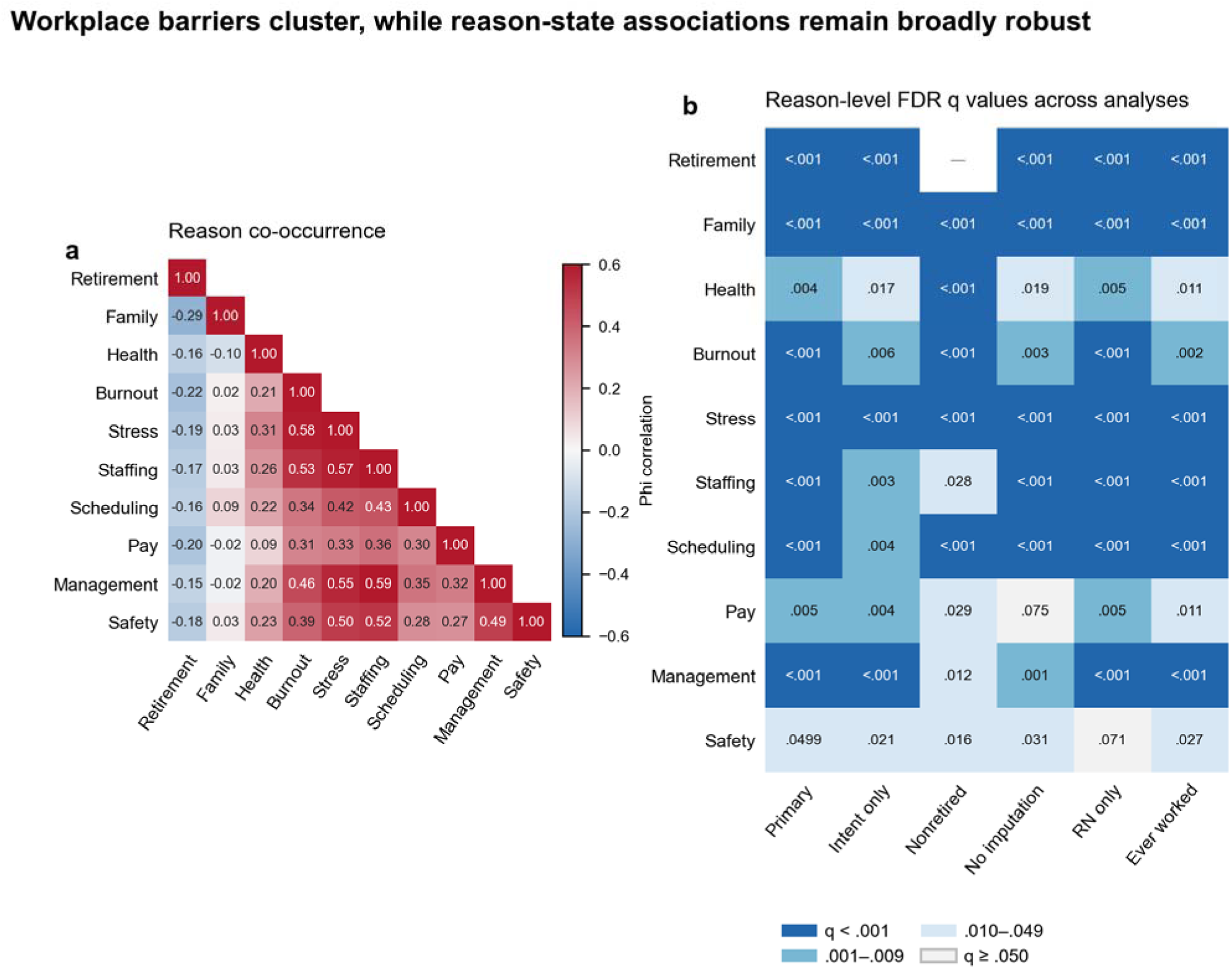
Workplace barriers cluster, while reason–state associations remain broadly robust. **a**, Survey-weighted pairwise phi correlations among the ten modeled reasons. Positive correlations show clustering of workplace barriers; retirement was inversely correlated with most workplace reasons. **b**, Reason-level Benjamini–Hochberg false-discovery-rate q values from finite-design joint F tests across the primary five-state analysis and five sensitivity analyses. Retirement was structurally omitted from the nonretirement analysis. The safety composite was attenuated in the RN-only analysis (q = 0.071), and low pay was attenuated after excluding records with an imputed outcome or reason block (q = 0.075).

### Retirement and family caregiving marked different reserves

Retirement had the clearest association with the no-future-intention state (Figure 2; Table 3). After standardization, respondents reporting retirement had a 60.1% probability of no future intention, compared with 33.1% among those not reporting retirement, a difference of 27.0 percentage points (95% CI 21.1 to 32.9; q < 0.001). Retirement was also associated with lower probabilities of active job search (−6.6 points, 95% CI −8.5 to −4.8), being undecided (−8.1 points, 95% CI −13.1 to −3.2), and already having entered or returned (−9.2 points, 95% CI −11.8 to −6.7). The reason-level joint test was significant (F4,79 = 37.77, q < 0.001).

**Table 3.** Adjusted marginal probability differences across workforce activation states. Cells are percentage-point differences (95% CI) for reason present versus absent from separate replicate-weight multinomial models. The last column is the Benjamini–Hochberg q value for the finite-design reason-level joint F test.

| Reason | No intention | Looking | Planned | Undecided | Entered/returned | Joint q |
| --- | --- | --- | --- | --- | --- | --- |
| Retirement | 27.0 (21.1 to 32.9) | -6.6 (-8.5 to -4.8) | -3.0 (-8.7 to 2.8) | -8.1 (-13.1 to -3.2) | -9.2 (-11.8 to -6.7) | <0.001 |
| Family caregiving | -13.3 (-18.0 to -8.5) | 1.3 (-1.4 to 4.0) | 14.6 (10.9 to 18.3) | 5.4 (1.4 to 9.5) | -8.0 (-10.7 to -5.4) | <0.001 |
| Disability, illness, or physical demands | 2.5 (-1.9 to 6.9) | -0.8 (-3.0 to 1.5) | -3.1 (-6.1 to -0.2) | 4.8 (0.7 to 8.8) | -3.4 (-6.4 to -0.3) | 0.004 |
| Burnout | 1.2 (-3.5 to 6.0) | -1.2 (-3.6 to 1.2) | -3.5 (-6.0 to -1.0) | 6.1 (1.6 to 10.6) | -2.6 (-5.1 to -0.2) | <0.001 |
| Stressful work environment | -2.1 (-6.7 to 2.5) | 0.1 (-2.4 to 2.5) | -4.2 (-6.7 to -1.7) | 8.5 (4.5 to 12.4) | -2.2 (-4.4 to -0.0) | <0.001 |
| Inadequate staffing | -3.9 (-9.9 to 2.2) | 0.6 (-2.3 to 3.5) | -2.3 (-4.9 to 0.3) | 9.8 (4.9 to 14.7) | -4.2 (-7.5 to -0.8) | <0.001 |
| Scheduling or hours | -9.2 (-15.8 to -2.5) | -0.6 (-3.6 to 2.3) | 0.9 (-2.9 to 4.6) | 14.6 (7.8 to 21.4) | -5.7 (-8.5 to -2.9) | <0.001 |
| Low pay | -0.1 (-6.2 to 6.0) | 1.5 (-1.8 to 4.9) | -4.9 (-7.7 to -2.1) | 6.4 (0.4 to 12.3) | -2.9 (-6.0 to 0.1) | 0.005 |
| Poor management or leadership | -7.1 (-11.9 to -2.3) | 2.7 (-0.5 to 5.9) | -3.7 (-6.3 to -1.1) | 9.6 (4.6 to 14.7) | -1.6 (-5.0 to 1.9) | <0.001 |
| Safety protocol, high-risk conditions, or workplace violence | -3.0 (-9.3 to 3.3) | 1.0 (-1.7 to 3.7) | -3.2 (-5.9 to -0.4) | 5.7 (0.4 to 11.1) | -0.6 (-3.7 to 2.6) | 0.050 |

Family caregiving showed the opposite pattern. Respondents reporting caregiving had a 21.2% adjusted probability of planned future entry or return, compared with 6.6% among those not reporting it, a difference of 14.6 points (95% CI 10.9 to 18.3; q < 0.001). Caregiving was associated with a 13.3-point lower probability of no future intention (95% CI −18.0 to −8.5) and an 8.0-point lower probability of already having entered or returned (95% CI −10.7 to −5.4). The probability of being undecided was 5.4 points higher (95% CI 1.4 to 9.5). The reason-level joint test was significant (F4,79 = 36.86, q < 0.001).

### Workplace barriers concentrated in the undecided state

Workplace barriers were consistently associated with a higher adjusted probability of being undecided (Figure 2; Table 3). The differences were 6.1 points for burnout (95% CI 1.6 to 10.6), 8.5 points for stressful work (95% CI 4.5 to 12.4), 9.8 points for inadequate staffing (95% CI 4.9 to 14.7), 14.6 points for scheduling or hours (95% CI 7.8 to 21.4), and 9.6 points for poor management or leadership (95% CI 4.6 to 14.7). Each of these contrasts remained significant after false-discovery-rate correction.

These reasons did not identify a pool already moving uniformly toward employment. Scheduling problems were associated with a lower probability of no future intention (−9.2 points, 95% CI −15.8 to −2.5) but also with a lower probability of already having entered or returned (−5.7 points, 95% CI −8.5 to −2.9). Inadequate staffing was associated with a 4.2-point lower probability of already having entered or returned (95% CI −7.5 to −0.8). Stress, burnout, low pay and poor management were each associated with lower probabilities of planned entry or return, although not every state-specific contrast survived multiplicity correction.

Health or physical limitations were associated with a 4.8-point higher probability of being undecided and a 3.4-point lower probability of already having entered or returned, but their individual contrast q values were near the 0.05 threshold. Low pay had a significant reason-level joint association in the primary analysis (F4,79 = 4.09, q = 0.006), driven mainly by a lower probability of planned entry or return. The safety composite had the weakest primary reason-level evidence (F4,79 = 2.49, q = 0.050); its higher undecided probability and lower planned-entry probability did not remain significant after correction across the 50 state-specific contrasts.

### Sensitivity analyses

The reason-state structure persisted across alternative specifications (Figure 3b; Supplementary Table S4). All ten reason-level tests remained significant when respondents who had already entered or returned were excluded. All nine nonretirement reasons remained significant after excluding respondents who selected retirement. Nine of ten remained significant after excluding officially imputed outcome and reason-block records; the association for low pay attenuated to q = 0.075. Nine of ten remained significant among non-APRN RNs; the safety composite attenuated to q = 0.071. All ten remained significant among respondents with previous nursing work or observed entry or return.

Official imputation influenced the estimated size of the observed-entry-or-return state but not the main ordering of reason profiles. The outcome was officially imputed for 805 of 7,753 target-sample respondents, and the entire reason block was imputed for 892. The weighted prevalence of already having entered or returned was 14.6% in the full target sample and approximately 11.4% when records with an imputed outcome or reason block were excluded.

## Discussion

This national analysis separates licensed capacity from workforce readiness. Among approximately 869,000 actively licensed RNs outside nursing employment, fewer than one in twenty were actively looking for nursing work and about one in seven had already entered or returned by the time of the survey. One quarter were undecided and nearly half had no future intention. The reasons for not working in nursing mapped to different positions within this distribution. Retirement identified a largely non-mobilizable group, family caregiving identified a planned reserve, and workplace barriers identified a conditional reserve whose members remained disproportionately undecided.

The findings extend previous NSSRN research in two ways. Castner and colleagues quantified the licensed nonworking RN pool using the 2018 NSSRN and identified willingness and return conditions as unresolved questions [7]. The official 2022 COVID brief distinguished return intentions among nurses who left during the pandemic [6]. Our analysis covers all actively licensed RNs outside nursing employment and separates active search, planned entry or return, uncertainty and observed entry or return. Recent NSSRN studies have clarified actual turnover among employed nurses [47–51]. The present estimates address activation from nonemployment, a distinct labor-market process.

Retirement accounted for much of the gap between licensure and mobilizable supply. Earlier US workforce projections established the importance of the RN age structure [40], and studies of retirement show that health, physical demands, income, flexibility and recognition shape extended working life [39]. At the population level, retirement was associated with little active search and a high probability of no future intention even after flexible age adjustment. Short-term supply models that count all licensed nonworkers as reserve capacity will consequently overstate the pool that recruitment campaigns can mobilize.

Family caregiving marked a different state. Caregivers were much more likely to plan a future entry or return and less likely to report no future intention, but they were also less likely to have already entered or returned. This pattern is consistent with a time-constrained planned reserve. Studies of inactive nurses and nurses with family-care responsibilities have repeatedly identified schedules, shorter shifts, part-time options and flexibility as relevant employment conditions [12,36,38]. Work-family conflict also has workplace antecedents and is associated with adverse employment outcomes [35,37]. These data do not evaluate a specific intervention, but they identify a population in which flexible scheduling, predictable hours and caregiving support can be tested as activation strategies.

Workplace barriers were concentrated in the undecided state rather than in active search or completed entry. The largest shift toward uncertainty occurred for scheduling or hours, followed by inadequate staffing, poor management and stressful work. This pattern helps reconcile two observations: nurses who report these barriers have not necessarily rejected nursing, yet they are not immediately available simply because vacancies exist. Staffing, shift length, burnout, leadership and work environment are established correlates of retention and turnover [19,23,25,28–30,52], and workplace violence adds a separate risk [32,33]. Our analysis locates these experiences after exit: they remain associated with the state in which a return decision has not been made. Recruitment incentives that leave workload, scheduling, management and safety unchanged may therefore reach the conditional reserve without converting it into active supply.

The co-occurrence results reinforce this interpretation. Burnout, stress, staffing, management and safety formed a connected work-environment cluster. This structure is consistent with evidence that burnout depends on workload and organizational context [26] and that supportive leadership and organizational commitment are central retention correlates [29,31]. The reason-specific models describe recognizable population profiles without partitioning the contribution of each correlated workplace exposure.

Pay and safety marked the boundary of the strongest evidence. Salary alone is not a complete account of nurse turnover [29], and the low-pay association attenuated after excluding officially imputed records. The safety composite combined high-risk conditions, safety protocols and violence; its reason-level evidence was borderline in the primary model and attenuated among non-APRN RNs. Prospective re-entry studies should measure these domains directly.

The results suggest a segmentation approach to workforce policy. First, supply forecasts should report licensed capacity separately from active search, planned entry or return, uncertainty and no intention. Second, outreach to the planned reserve should be paired with practical supports for caregiving and scheduling. Third, activation of the conditional reserve requires changes to the employment offer itself, including manageable staffing, predictable schedules, credible leadership and workplace safety. Fourth, re-entry programs should distinguish people who need skills updating or placement assistance from those whose central barrier is work quality. Evidence for specific turnover-reduction interventions remains less developed than the observational literature [53], so these strategies require prospective evaluation using actual entry and sustained employment rather than intention alone.

This study has several strengths. It uses a large nationally representative survey designed specifically for the US nursing workforce, preserves five substantively different activation states, incorporates all official replicate weights, and reports marginal probability differences that are directly interpretable for policy. The inclusion of all 23 official reasons prevents selective description, while minimum cell-size rules and prespecified reason domains limit unstable inference. Sensitivity analyses address retirement, APRN status, official imputation and the presence of respondents who had never worked in nursing.

The study has several limitations. The cross-sectional design cannot establish that a reported reason caused an activation state. Respondents could select multiple reasons, and workplace reasons were strongly correlated. Those who had already entered or returned did so after 31 December 2021, but the public-use file does not provide a common follow-up interval for estimating a return rate. Official single imputation was used for approximately 10% of outcomes and 11.5% of reason blocks; replicate weights do not propagate imputation uncertainty. The smallest activation state contained 292 respondents, preventing adjusted analysis of uncommon reasons. Finally, 774 target-sample respondents had never worked in nursing, so the main estimand concerns entry or re-entry activation rather than return alone. The strict ever-worked sensitivity analysis produced the same reason-level structure.

## Conclusions

Licensed RNs outside nursing employment form a heterogeneous reserve. Retirement, family caregiving and workplace barriers identify populations at different distances from effective labor supply. Measuring activation state converts a nominal count of licensed capacity into a more realistic account of who is unavailable, who plans to enter or return, and whose decision remains conditional on the quality and organization of nursing work.

## Declarations

### Ethics approval and consent to participate

In accordance with the authors’ institutional policy, this secondary analysis of the publicly available de-identified 2022 NSSRN public-use file did not require local ethics committee review. The investigators had no interaction with participants, no access to direct identifiers and made no attempt to identify respondents. Participation in the original NSSRN was voluntary. The US Census Bureau protected responses under Title 13, United States Code, section 9; the federal information collection was approved under OMB Control No. 0607-1002. Additional informed consent was not required for this secondary analysis.

## Consent for publication

Not applicable.

## Data availability

The 2022 NSSRN public-use file, questionnaire and documentation are available from the HRSA Health Workforce data portal. The analysis code and aggregate source data underlying every figure and table accompany this submission. The study does not redistribute respondent-level data.

## Code availability

The complete R analysis code used to define the study population, fit replicate-weight multinomial models and generate tables is included in the submission package. Python code used to generate the publication figures is also included.

## Author contributions

MH and ZC conceived and designed the study. MH curated the data, performed the statistical analysis, and drafted the original manuscript. ZC supervised the study, contributed to methodology and interpretation, and critically revised the manuscript. Both authors read and approved the final manuscript.

## Acknowledgements

The authors acknowledge the Health Resources and Services Administration, National Center for Health Workforce Analysis, and the US Census Bureau for developing the 2022 National Sample Survey of Registered Nurses and making the public-use data available. The findings and conclusions are those of the authors and do not necessarily represent the views of HRSA or the US Census Bureau.

## Artificial intelligence use

During preparation of this work, the authors used OpenAI Codex to assist with code generation and language editing. The authors reviewed and verified all analytic code and text, revised the outputs where necessary, and take full responsibility for the integrity, accuracy and conclusions of the manuscript.

