## Supplementary_Material for "Distinct workforce activation states among licensed US nurses outside nursing employment"

### Supplementary methods

The analytic population comprised respondents in the 2022 NSSRN public-use file who held an RN or RN–APRN license and were not working for pay in nursing on 31 December 2021. The replicate-weight survey design was created before restricting to this population, so all estimates used domain analysis. The full model sample excluded nine respondents with missing highest nursing education.

The primary outcome retained five mutually exclusive employment-activation states. Ten modeled reasons met the prespecified minimum of 30 endorsers in every reason-by-state cell. Health combined disability or illness with physical demands; safety combined high-risk working conditions, unsatisfactory safety protocol and workplace harassment or violence. Each reason was analyzed in a separate multinomial model because multiple reasons could be selected and workplace reasons clustered strongly.

Variance estimation used the final person weight and 80 successive-difference replicate weights. The replicate variance was (4/80) multiplied by the sum of squared differences between each replicate estimate and the full-sample estimate. Reason-level four-degree-of-freedom Wald statistics were evaluated as F tests with 79 denominator degrees of freedom. Benjamini–Hochberg correction was applied separately to the ten joint tests and 50 marginal contrasts.

The complete machine-readable versions of every table are supplied in the supplementary_tables directory. Table S1 characteristics are also supplied there as a long-format numeric file for reuse.

### STROBE checklist

| **Item** | **Recommendation** | **How addressed** | **Location** |
| --- | --- | --- | --- |
| 1 | Title and abstract | Design identified in title/abstract; structured summary reports population, methods and principal estimates. | Title; Abstract |
| 2 | Background/rationale | Scientific background and workforce-planning rationale provided. | Introduction |
| 3 | Objectives | Three study questions and prespecified expectations stated. | Introduction, final paragraph |
| 4 | Study design | National cross-sectional secondary survey analysis described early. | Methods—Study design and data source |
| 5 | Setting | Survey sponsor, administrator, dates, US coverage and reference date reported. | Methods—Study design and data source |
| 6 | Participants | Eligibility, exclusions and model-eligible sample reported. | Methods—Study population |
| 7 | Variables | Five activation states, 23 reasons, modeled domains and covariates defined. | Methods—Outcome, reasons and covariates |
| 8 | Data sources/measurement | Official public-use variables, weights and composite reason definitions described. | Methods |
| 9 | Bias | Domain analysis, complete covariates, official imputation and sensitivity analyses specified. | Methods—Statistical analysis |
| 10 | Study size | Census of eligible survey respondents; minimum cell rule for modeled reasons reported. | Methods—Study population and reasons |
| 11 | Quantitative variables | Age spline and categorical grouping decisions reported. | Methods—Covariates |
| 12a | Statistical methods | Replicate-weight multinomial models, standardization, joint tests and FDR correction reported. | Methods—Statistical analysis |
| 12b | Subgroups/interactions | No interaction analysis; five prespecified sensitivity restrictions reported. | Methods—Statistical analysis |
| 12c | Missing data | Nine covariate exclusions and official imputation sensitivity analysis reported. | Methods; Results—Sensitivity analyses |
| 12d | Sampling strategy | Main weight, 80 SDR weights, variance formula and domain analysis reported. | Methods—Statistical analysis |
| 12e | Sensitivity analyses | Five alternative population/imputation definitions reported. | Methods; Results |
| 13 | Participants | Counts at target and model stages and exclusion reason reported. | Methods—Study population; Results |
| 14 | Descriptive data | Weighted demographic, professional and household characteristics reported. | Results; Table 1 |
| 15 | Outcome data | Counts and weighted prevalence for all five states reported. | Results; Figure 1 |
| 16 | Main results | Adjusted standardized differences, 95% CIs, q values and covariates reported. | Results; Tables 2–3; Figure 2 |
| 17 | Other analyses | Correlation structure, imputation comparison and five sensitivity analyses reported. | Results; Figure 3; Supplement |
| 18 | Key results | Findings summarized against objectives without causal claims. | Discussion, opening paragraph |
| 19 | Limitations | Direction and scope of major design and measurement limitations discussed. | Discussion—Limitations paragraph |
| 20 | Interpretation | Evidence, prior literature and policy implications integrated cautiously. | Discussion |
| 21 | Generalisability | National target population and limits of the public-use estimand described. | Methods; Discussion |
| 22 | Funding | No specific funding reported; funder role not applicable. | Declarations—Funding |

**Supplementary Table S1 | Activation-state distribution among respondents endorsing each reason**

Percentages are calculated among endorsers of each reason and use survey weights with 95% confidence intervals. The sparse-cell flag is based on unweighted n < 30.

| **Reason** | **Activation state** | **n** | **Weighted N** | **% (95% CI)** | **n <30** |
| --- | --- | --- | --- | --- | --- |
| Retirement | No future intention | 2,737 | 257,574 | 70.0 (67.4 to 72.6) | FALSE |
| Retirement | Actively looking | 44 | 1,765 | 0.5 (0.2 to 0.8) | FALSE |
| Retirement | Plans future entry or return | 108 | 13,224 | 3.6 (2.5 to 4.7) | FALSE |
| Retirement | Undecided | 977 | 88,523 | 24.1 (21.7 to 26.4) | FALSE |
| Retirement | Already entered or returned | 122 | 6,929 | 1.9 (1.4 to 2.3) | FALSE |
| Family caregiving | No future intention | 368 | 45,378 | 23.5 (19.8 to 27.2) | FALSE |
| Family caregiving | Actively looking | 93 | 12,862 | 6.7 (4.0 to 9.3) | FALSE |
| Family caregiving | Plans future entry or return | 372 | 52,286 | 27.1 (23.1 to 31.1) | FALSE |
| Family caregiving | Undecided | 515 | 54,581 | 28.3 (24.9 to 31.7) | FALSE |
| Family caregiving | Already entered or returned | 223 | 27,797 | 14.4 (11.5 to 17.3) | FALSE |
| Disability or illness | No future intention | 342 | 41,655 | 44.0 (38.7 to 49.2) | FALSE |
| Disability or illness | Actively looking | 41 | 5,440 | 5.7 (2.4 to 9.1) | FALSE |
| Disability or illness | Plans future entry or return | 91 | 9,489 | 10.0 (6.6 to 13.5) | FALSE |
| Disability or illness | Undecided | 247 | 29,719 | 31.4 (26.5 to 36.3) | FALSE |
| Disability or illness | Already entered or returned | 58 | 8,387 | 8.9 (6.2 to 11.5) | FALSE |
| COVID-19 severe-illness risk condition (self/family) | No future intention | 130 | 15,255 | 31.0 (23.8 to 38.1) | FALSE |
| COVID-19 severe-illness risk condition (self/family) | Actively looking | 22 | 2,313 | 4.7 (0.0 to 10.1) | TRUE |
| COVID-19 severe-illness risk condition (self/family) | Plans future entry or return | 59 | 7,360 | 14.9 (8.8 to 21.1) | FALSE |
| COVID-19 severe-illness risk condition (self/family) | Undecided | 143 | 15,506 | 31.5 (23.4 to 39.5) | FALSE |
| COVID-19 severe-illness risk condition (self/family) | Already entered or returned | 64 | 8,834 | 17.9 (10.3 to 25.6) | FALSE |
| Physical demands of the job | No future intention | 307 | 42,511 | 42.4 (36.5 to 48.3) | FALSE |
| Physical demands of the job | Actively looking | 29 | 3,878 | 3.9 (1.3 to 6.4) | TRUE |
| Physical demands of the job | Plans future entry or return | 50 | 7,258 | 7.2 (4.1 to 10.4) | FALSE |
| Physical demands of the job | Undecided | 234 | 31,741 | 31.7 (26.2 to 37.2) | FALSE |
| Physical demands of the job | Already entered or returned | 101 | 14,871 | 14.8 (11.3 to 18.3) | FALSE |
| Burnout | No future intention | 533 | 59,149 | 33.6 (29.3 to 37.8) | FALSE |
| Burnout | Actively looking | 72 | 9,286 | 5.3 (2.4 to 8.1) | FALSE |
| Burnout | Plans future entry or return | 163 | 20,652 | 11.7 (8.4 to 15.0) | FALSE |
| Burnout | Undecided | 435 | 51,777 | 29.4 (25.1 to 33.7) | FALSE |
| Burnout | Already entered or returned | 272 | 35,418 | 20.1 (16.4 to 23.8) | FALSE |
| Stressful work environment | No future intention | 469 | 52,584 | 33.3 (29.5 to 37.1) | FALSE |
| Stressful work environment | Actively looking | 63 | 9,536 | 6.0 (3.1 to 8.9) | FALSE |
| Stressful work environment | Plans future entry or return | 123 | 15,734 | 10.0 (7.1 to 12.8) | FALSE |
| Stressful work environment | Undecided | 419 | 49,823 | 31.5 (27.8 to 35.3) | FALSE |
| Stressful work environment | Already entered or returned | 213 | 30,255 | 19.2 (15.9 to 22.4) | FALSE |
| Inadequate staffing | No future intention | 315 | 43,268 | 31.8 (26.4 to 37.1) | FALSE |
| Inadequate staffing | Actively looking | 41 | 9,515 | 7.0 (3.2 to 10.8) | FALSE |
| Inadequate staffing | Plans future entry or return | 103 | 16,479 | 12.1 (8.8 to 15.4) | FALSE |
| Inadequate staffing | Undecided | 318 | 43,958 | 32.3 (27.5 to 37.0) | FALSE |
| Inadequate staffing | Already entered or returned | 147 | 23,016 | 16.9 (12.3 to 21.5) | FALSE |
| Scheduling or hours | No future intention | 207 | 26,759 | 27.6 (22.6 to 32.6) | FALSE |
| Scheduling or hours | Actively looking | 42 | 5,124 | 5.3 (1.8 to 8.7) | FALSE |
| Scheduling or hours | Plans future entry or return | 117 | 15,227 | 15.7 (11.2 to 20.2) | FALSE |
| Scheduling or hours | Undecided | 270 | 35,592 | 36.7 (30.3 to 43.1) | FALSE |
| Scheduling or hours | Already entered or returned | 119 | 14,288 | 14.7 (10.8 to 18.7) | FALSE |
| Low pay or better pay elsewhere | No future intention | 199 | 24,502 | 29.2 (23.2 to 35.3) | FALSE |
| Low pay or better pay elsewhere | Actively looking | 45 | 7,830 | 9.3 (4.7 to 14.0) | FALSE |
| Low pay or better pay elsewhere | Plans future entry or return | 72 | 8,768 | 10.5 (6.3 to 14.6) | FALSE |
| Low pay or better pay elsewhere | Undecided | 182 | 24,667 | 29.4 (23.8 to 35.1) | FALSE |
| Low pay or better pay elsewhere | Already entered or returned | 116 | 18,077 | 21.6 (15.7 to 27.4) | FALSE |
| Poor management or leadership | No future intention | 322 | 33,679 | 29.3 (24.7 to 33.9) | FALSE |
| Poor management or leadership | Actively looking | 57 | 10,313 | 9.0 (4.7 to 13.2) | FALSE |
| Poor management or leadership | Plans future entry or return | 83 | 11,747 | 10.2 (6.7 to 13.8) | FALSE |
| Poor management or leadership | Undecided | 313 | 36,751 | 32.0 (27.0 to 37.0) | FALSE |
| Poor management or leadership | Already entered or returned | 168 | 22,415 | 19.5 (14.5 to 24.5) | FALSE |
| Poor collaboration or communication | No future intention | 99 | 12,907 | 32.6 (23.4 to 41.7) | FALSE |
| Poor collaboration or communication | Actively looking | 25 | 4,461 | 11.3 (3.0 to 19.5) | TRUE |
| Poor collaboration or communication | Plans future entry or return | 31 | 2,979 | 7.5 (3.3 to 11.7) | FALSE |
| Poor collaboration or communication | Undecided | 122 | 14,556 | 36.7 (27.7 to 45.7) | FALSE |
| Poor collaboration or communication | Already entered or returned | 44 | 4,736 | 11.9 (5.2 to 18.7) | FALSE |
| Lack of advancement opportunities | No future intention | 68 | 7,535 | 27.6 (17.5 to 37.8) | FALSE |
| Lack of advancement opportunities | Actively looking | 17 | 3,785 | 13.9 (3.5 to 24.3) | TRUE |
| Lack of advancement opportunities | Plans future entry or return | 24 | 3,247 | 11.9 (6.1 to 17.7) | TRUE |
| Lack of advancement opportunities | Undecided | 71 | 8,129 | 29.8 (18.2 to 41.5) | FALSE |
| Lack of advancement opportunities | Already entered or returned | 36 | 4,555 | 16.7 (9.3 to 24.1) | FALSE |
| High-risk working conditions | No future intention | 216 | 28,697 | 31.2 (25.8 to 36.7) | FALSE |
| High-risk working conditions | Actively looking | 38 | 5,858 | 6.4 (2.9 to 9.9) | FALSE |
| High-risk working conditions | Plans future entry or return | 64 | 8,813 | 9.6 (6.1 to 13.1) | FALSE |
| High-risk working conditions | Undecided | 245 | 28,176 | 30.7 (24.9 to 36.4) | FALSE |
| High-risk working conditions | Already entered or returned | 123 | 20,305 | 22.1 (17.1 to 27.1) | FALSE |
| Unsatisfactory safety protocol | No future intention | 118 | 14,934 | 28.0 (19.6 to 36.3) | FALSE |
| Unsatisfactory safety protocol | Actively looking | 27 | 4,485 | 8.4 (3.3 to 13.5) | TRUE |
| Unsatisfactory safety protocol | Plans future entry or return | 47 | 7,653 | 14.3 (8.1 to 20.6) | FALSE |
| Unsatisfactory safety protocol | Undecided | 123 | 14,851 | 27.8 (20.4 to 35.2) | FALSE |
| Unsatisfactory safety protocol | Already entered or returned | 70 | 11,466 | 21.5 (13.8 to 29.2) | FALSE |
| Workplace harassment or violence | No future intention | 67 | 10,080 | 29.7 (17.7 to 41.7) | FALSE |
| Workplace harassment or violence | Actively looking | 15 | 1,913 | 5.6 (0.0 to 12.7) | TRUE |
| Workplace harassment or violence | Plans future entry or return | 31 | 4,794 | 14.1 (5.5 to 22.7) | FALSE |
| Workplace harassment or violence | Undecided | 106 | 11,355 | 33.4 (22.8 to 44.0) | FALSE |
| Workplace harassment or violence | Already entered or returned | 53 | 5,810 | 17.1 (9.7 to 24.6) | FALSE |
| Liability concerns | No future intention | 124 | 17,463 | 32.4 (23.7 to 41.2) | FALSE |
| Liability concerns | Actively looking | 26 | 4,147 | 7.7 (1.2 to 14.2) | TRUE |
| Liability concerns | Plans future entry or return | 43 | 6,624 | 12.3 (7.1 to 17.5) | FALSE |
| Liability concerns | Undecided | 146 | 15,798 | 29.3 (22.2 to 36.5) | FALSE |
| Liability concerns | Already entered or returned | 63 | 9,847 | 18.3 (11.7 to 24.9) | FALSE |
| Inability to practice nursing professionally | No future intention | 52 | 3,558 | 18.7 (10.8 to 26.5) | FALSE |
| Inability to practice nursing professionally | Actively looking | 19 | 3,101 | 16.3 (2.0 to 30.5) | TRUE |
| Inability to practice nursing professionally | Plans future entry or return | 9 | 518 | 2.7 (0.0 to 5.7) | TRUE |
| Inability to practice nursing professionally | Undecided | 50 | 5,733 | 30.1 (20.0 to 40.1) | FALSE |
| Inability to practice nursing professionally | Already entered or returned | 30 | 6,165 | 32.3 (16.3 to 48.4) | FALSE |
| Career change | No future intention | 355 | 52,294 | 48.0 (42.8 to 53.2) | FALSE |
| Career change | Actively looking | 20 | 4,976 | 4.6 (1.8 to 7.4) | TRUE |
| Career change | Plans future entry or return | 59 | 8,464 | 7.8 (5.1 to 10.4) | FALSE |
| Career change | Undecided | 237 | 33,178 | 30.5 (24.5 to 36.4) | FALSE |
| Career change | Already entered or returned | 92 | 10,046 | 9.2 (6.4 to 12.1) | FALSE |
| School or educational program | No future intention | 21 | 3,619 | 10.0 (4.4 to 15.5) | TRUE |
| School or educational program | Actively looking | 22 | 3,797 | 10.5 (3.3 to 17.7) | TRUE |
| School or educational program | Plans future entry or return | 55 | 10,428 | 28.8 (16.5 to 41.0) | FALSE |
| School or educational program | Undecided | 11 | 889 | 2.5 (0.3 to 4.6) | TRUE |
| School or educational program | Already entered or returned | 102 | 17,522 | 48.3 (36.9 to 59.7) | FALSE |
| Skills are out of date | No future intention | 114 | 13,085 | 42.1 (33.6 to 50.6) | FALSE |
| Skills are out of date | Actively looking | 15 | 1,379 | 4.4 (0.0 to 9.6) | TRUE |
| Skills are out of date | Plans future entry or return | 19 | 1,648 | 5.3 (1.6 to 9.0) | TRUE |
| Skills are out of date | Undecided | 110 | 13,187 | 42.5 (33.6 to 51.3) | FALSE |
| Skills are out of date | Already entered or returned | 17 | 1,763 | 5.7 (1.2 to 10.1) | TRUE |
| Difficulty finding a nursing position | No future intention | 30 | 2,873 | 10.8 (3.3 to 18.2) | FALSE |
| Difficulty finding a nursing position | Actively looking | 51 | 8,367 | 31.3 (13.7 to 49.0) | FALSE |
| Difficulty finding a nursing position | Plans future entry or return | 27 | 2,512 | 9.4 (2.1 to 16.7) | TRUE |
| Difficulty finding a nursing position | Undecided | 61 | 4,594 | 17.2 (8.3 to 26.2) | FALSE |
| Difficulty finding a nursing position | Already entered or returned | 77 | 8,349 | 31.3 (17.9 to 44.7) | FALSE |
| Other | No future intention | 81 | 8,297 | 15.6 (10.4 to 20.9) | FALSE |
| Other | Actively looking | 20 | 2,871 | 5.4 (2.2 to 8.6) | TRUE |
| Other | Plans future entry or return | 29 | 2,965 | 5.6 (2.1 to 9.0) | TRUE |
| Other | Undecided | 91 | 9,846 | 18.5 (11.3 to 25.8) | FALSE |
| Other | Already entered or returned | 198 | 29,139 | 54.9 (45.7 to 64.0) | FALSE |

**Supplementary Table S2 | Primary replicate-weight multinomial relative risk ratios**

The reference outcome is no future intention. Separate models were fitted for each reason. All models adjusted for age, sex, race and ethnicity, nursing education and license type; the caregiving model also adjusted for marital status and dependents.

| **Reason** | **Outcome vs no intention** | **RRR (95% CI)** | **p** | **Contrast q** | **Joint q** |
| --- | --- | --- | --- | --- | --- |
| Burnout | Actively looking | 0.64 (0.32 to 1.30) | 0.216 | 0.340 | <0.001 |
| Burnout | Plans future entry or return | 0.59 (0.40 to 0.86) | 0.007 | 0.025 | <0.001 |
| Burnout | Undecided | 1.18 (0.91 to 1.53) | 0.221 | 0.340 | <0.001 |
| Burnout | Already entered or returned | 0.67 (0.50 to 0.91) | 0.010 | 0.030 | <0.001 |
| Family caregiving | Actively looking | 2.02 (1.07 to 3.82) | 0.030 | 0.067 | <0.001 |
| Family caregiving | Plans future entry or return | 5.02 (3.56 to 7.09) | <0.001 | <0.001 | <0.001 |
| Family caregiving | Undecided | 1.77 (1.38 to 2.28) | <0.001 | <0.001 | <0.001 |
| Family caregiving | Already entered or returned | 0.85 (0.57 to 1.27) | 0.430 | 0.538 | <0.001 |
| Disability, illness, or physical demands | Actively looking | 0.70 (0.37 to 1.32) | 0.264 | 0.377 | 0.004 |
| Disability, illness, or physical demands | Plans future entry or return | 0.59 (0.41 to 0.87) | 0.007 | 0.025 | 0.004 |
| Disability, illness, or physical demands | Undecided | 1.09 (0.86 to 1.37) | 0.489 | 0.575 | 0.004 |
| Disability, illness, or physical demands | Already entered or returned | 0.62 (0.42 to 0.90) | 0.013 | 0.038 | 0.004 |
| Poor management or leadership | Actively looking | 1.97 (1.10 to 3.53) | 0.022 | 0.051 | <0.001 |
| Poor management or leadership | Plans future entry or return | 0.79 (0.51 to 1.24) | 0.307 | 0.401 | <0.001 |
| Poor management or leadership | Undecided | 1.67 (1.28 to 2.18) | <0.001 | <0.001 | <0.001 |
| Poor management or leadership | Already entered or returned | 1.05 (0.69 to 1.61) | 0.818 | 0.884 | <0.001 |
| Low pay | Actively looking | 1.23 (0.60 to 2.49) | 0.571 | 0.653 | 0.005 |
| Low pay | Plans future entry or return | 0.51 (0.29 to 0.89) | 0.019 | 0.047 | 0.005 |
| Low pay | Undecided | 1.23 (0.89 to 1.70) | 0.218 | 0.340 | 0.005 |
| Low pay | Already entered or returned | 0.70 (0.45 to 1.08) | 0.105 | 0.210 | 0.005 |
| Retirement | Actively looking | 0.04 (0.02 to 0.09) | <0.001 | <0.001 | <0.001 |
| Retirement | Plans future entry or return | 0.33 (0.16 to 0.64) | 0.001 | 0.006 | <0.001 |
| Retirement | Undecided | 0.39 (0.30 to 0.51) | <0.001 | <0.001 | <0.001 |
| Retirement | Already entered or returned | 0.18 (0.12 to 0.25) | <0.001 | <0.001 | <0.001 |
| Safety protocol, high-risk conditions, or workplace violence | Actively looking | 1.28 (0.66 to 2.49) | 0.459 | 0.556 | 0.050 |
| Safety protocol, high-risk conditions, or workplace violence | Plans future entry or return | 0.75 (0.47 to 1.21) | 0.237 | 0.351 | 0.050 |
| Safety protocol, high-risk conditions, or workplace violence | Undecided | 1.32 (0.95 to 1.83) | 0.098 | 0.206 | 0.050 |
| Safety protocol, high-risk conditions, or workplace violence | Already entered or returned | 1.00 (0.66 to 1.50) | 0.988 | 0.988 | 0.050 |
| Scheduling or hours | Actively looking | 1.03 (0.45 to 2.36) | 0.945 | 0.969 | <0.001 |
| Scheduling or hours | Plans future entry or return | 1.29 (0.80 to 2.08) | 0.297 | 0.401 | <0.001 |
| Scheduling or hours | Undecided | 2.02 (1.40 to 2.92) | <0.001 | <0.001 | <0.001 |
| Scheduling or hours | Already entered or returned | 0.74 (0.47 to 1.14) | 0.173 | 0.302 | <0.001 |
| Inadequate staffing | Actively looking | 1.15 (0.56 to 2.35) | 0.712 | 0.791 | <0.001 |
| Inadequate staffing | Plans future entry or return | 0.79 (0.50 to 1.24) | 0.311 | 0.401 | <0.001 |
| Inadequate staffing | Undecided | 1.52 (1.13 to 2.05) | 0.006 | 0.025 | <0.001 |
| Inadequate staffing | Already entered or returned | 0.71 (0.44 to 1.14) | 0.157 | 0.285 | <0.001 |
| Stressful work environment | Actively looking | 0.97 (0.52 to 1.80) | 0.924 | 0.969 | <0.001 |
| Stressful work environment | Plans future entry or return | 0.60 (0.40 to 0.92) | 0.018 | 0.047 | <0.001 |
| Stressful work environment | Undecided | 1.39 (1.09 to 1.77) | 0.007 | 0.025 | <0.001 |
| Stressful work environment | Already entered or returned | 0.79 (0.59 to 1.06) | 0.110 | 0.210 | <0.001 |

**Supplementary Table S3 | Reason-level joint tests across the primary and sensitivity analyses**

Finite-design F tests use 79 denominator degrees of freedom. q values are Benjamini–Hochberg adjusted within analysis.

| **Analysis** | **Reason** | **F** | **df** | **p** | **q** |
| --- | --- | --- | --- | --- | --- |
| Primary | Retirement | 37.77 | 4, 79 | <0.001 | <0.001 |
| Primary | Family caregiving | 36.86 | 4, 79 | <0.001 | <0.001 |
| Primary | Disability, illness, or physical demands | 4.29 | 4, 79 | 0.003 | 0.004 |
| Primary | Burnout | 5.98 | 4, 79 | <0.001 | <0.001 |
| Primary | Stressful work environment | 8.56 | 4, 79 | <0.001 | <0.001 |
| Primary | Inadequate staffing | 5.46 | 4, 79 | <0.001 | <0.001 |
| Primary | Scheduling or hours | 8.78 | 4, 79 | <0.001 | <0.001 |
| Primary | Low pay | 4.09 | 4, 79 | 0.005 | 0.005 |
| Primary | Poor management or leadership | 6.98 | 4, 79 | <0.001 | <0.001 |
| Primary | Safety protocol, high-risk conditions, or workplace violence | 2.49 | 4, 79 | 0.050 | 0.050 |
| Intent only | Retirement | 31.09 | 3, 79 | <0.001 | <0.001 |
| Intent only | Family caregiving | 28.72 | 3, 79 | <0.001 | <0.001 |
| Intent only | Disability, illness, or physical demands | 3.67 | 3, 79 | 0.016 | 0.017 |
| Intent only | Burnout | 4.63 | 3, 79 | 0.005 | 0.006 |
| Intent only | Stressful work environment | 8.43 | 3, 79 | <0.001 | <0.001 |
| Intent only | Inadequate staffing | 5.54 | 3, 79 | 0.002 | 0.003 |
| Intent only | Scheduling or hours | 5.33 | 3, 79 | 0.002 | 0.004 |
| Intent only | Low pay | 5.05 | 3, 79 | 0.003 | 0.004 |
| Intent only | Poor management or leadership | 9.45 | 3, 79 | <0.001 | <0.001 |
| Intent only | Safety protocol, high-risk conditions, or workplace violence | 3.44 | 3, 79 | 0.021 | 0.021 |
| Nonretired | Family caregiving | 30.63 | 4, 79 | <0.001 | <0.001 |
| Nonretired | Disability, illness, or physical demands | 8.65 | 4, 79 | <0.001 | <0.001 |
| Nonretired | Burnout | 7.26 | 4, 79 | <0.001 | <0.001 |
| Nonretired | Stressful work environment | 9.18 | 4, 79 | <0.001 | <0.001 |
| Nonretired | Inadequate staffing | 2.95 | 4, 79 | 0.025 | 0.028 |
| Nonretired | Scheduling or hours | 6.22 | 4, 79 | <0.001 | <0.001 |
| Nonretired | Low pay | 2.85 | 4, 79 | 0.029 | 0.029 |
| Nonretired | Poor management or leadership | 3.74 | 4, 79 | 0.008 | 0.012 |
| Nonretired | Safety protocol, high-risk conditions, or workplace violence | 3.41 | 4, 79 | 0.013 | 0.016 |
| No imputation | Retirement | 25.62 | 4, 79 | <0.001 | <0.001 |
| No imputation | Family caregiving | 21.50 | 4, 79 | <0.001 | <0.001 |
| No imputation | Disability, illness, or physical demands | 3.29 | 4, 79 | 0.015 | 0.019 |
| No imputation | Burnout | 4.74 | 4, 79 | 0.002 | 0.003 |
| No imputation | Stressful work environment | 5.83 | 4, 79 | <0.001 | <0.001 |
| No imputation | Inadequate staffing | 5.70 | 4, 79 | <0.001 | <0.001 |
| No imputation | Scheduling or hours | 5.77 | 4, 79 | <0.001 | <0.001 |
| No imputation | Low pay | 2.21 | 4, 79 | 0.075 | 0.075 |
| No imputation | Poor management or leadership | 5.21 | 4, 79 | <0.001 | 0.001 |
| No imputation | Safety protocol, high-risk conditions, or workplace violence | 2.88 | 4, 79 | 0.028 | 0.031 |
| RN only | Retirement | 32.05 | 4, 79 | <0.001 | <0.001 |
| RN only | Family caregiving | 33.82 | 4, 79 | <0.001 | <0.001 |
| RN only | Disability, illness, or physical demands | 4.17 | 4, 79 | 0.004 | 0.005 |
| RN only | Burnout | 5.47 | 4, 79 | <0.001 | <0.001 |
| RN only | Stressful work environment | 7.14 | 4, 79 | <0.001 | <0.001 |
| RN only | Inadequate staffing | 5.50 | 4, 79 | <0.001 | <0.001 |
| RN only | Scheduling or hours | 8.65 | 4, 79 | <0.001 | <0.001 |
| RN only | Low pay | 4.18 | 4, 79 | 0.004 | 0.005 |
| RN only | Poor management or leadership | 6.61 | 4, 79 | <0.001 | <0.001 |
| RN only | Safety protocol, high-risk conditions, or workplace violence | 2.25 | 4, 79 | 0.071 | 0.071 |
| Ever worked | Retirement | 38.51 | 4, 79 | <0.001 | <0.001 |
| Ever worked | Family caregiving | 30.17 | 4, 79 | <0.001 | <0.001 |
| Ever worked | Disability, illness, or physical demands | 3.62 | 4, 79 | 0.009 | 0.011 |
| Ever worked | Burnout | 5.05 | 4, 79 | 0.001 | 0.002 |
| Ever worked | Stressful work environment | 9.22 | 4, 79 | <0.001 | <0.001 |
| Ever worked | Inadequate staffing | 8.06 | 4, 79 | <0.001 | <0.001 |
| Ever worked | Scheduling or hours | 9.99 | 4, 79 | <0.001 | <0.001 |
| Ever worked | Low pay | 3.60 | 4, 79 | 0.010 | 0.011 |
| Ever worked | Poor management or leadership | 9.29 | 4, 79 | <0.001 | <0.001 |
| Ever worked | Safety protocol, high-risk conditions, or workplace violence | 2.90 | 4, 79 | 0.027 | 0.027 |

**Supplementary Table S4 | Comparison of all formal-model records with the nonimputed sensitivity sample**

The difference column is the percentage-point difference from the full formal-model sample.

| **Measure** | **Item** | **Sample** | **n** | **% (95% CI)** | **Difference, pp** |
| --- | --- | --- | --- | --- | --- |
| Activation state | No future intention | All formal-model records | 3,696 | 44.4 (42.7 to 46.2) | +0.0 |
| Activation state | No future intention | Outcome not imputed | 3,409 | 46.0 (44.0 to 48.0) | +1.6 |
| Activation state | No future intention | Reason block not imputed | 3,430 | 47.3 (45.2 to 49.4) | +2.9 |
| Activation state | No future intention | Outcome and reason block not imputed | 3,374 | 47.3 (45.3 to 49.4) | +2.9 |
| Activation state | Actively looking | All formal-model records | 292 | 4.7 (3.6 to 5.8) | +0.0 |
| Activation state | Actively looking | Outcome not imputed | 256 | 4.1 (3.1 to 5.0) | -0.6 |
| Activation state | Actively looking | Reason block not imputed | 244 | 4.1 (3.1 to 5.1) | -0.6 |
| Activation state | Actively looking | Outcome and reason block not imputed | 242 | 4.1 (3.1 to 5.1) | -0.6 |
| Activation state | Plans future entry or return | Outcome and reason block not imputed | 596 | 10.5 (9.0 to 12.0) | -0.2 |
| Activation state | Plans future entry or return | Outcome not imputed | 602 | 10.2 (8.7 to 11.6) | -0.5 |
| Activation state | Plans future entry or return | All formal-model records | 697 | 10.7 (9.3 to 12.1) | +0.0 |
| Activation state | Plans future entry or return | Reason block not imputed | 601 | 10.4 (8.9 to 11.9) | -0.3 |
| Activation state | Undecided | Outcome not imputed | 1,861 | 26.1 (24.3 to 27.9) | +0.5 |
| Activation state | Undecided | All formal-model records | 2,086 | 25.6 (24.0 to 27.2) | +0.0 |
| Activation state | Undecided | Outcome and reason block not imputed | 1,830 | 26.7 (24.9 to 28.5) | +1.1 |
| Activation state | Undecided | Reason block not imputed | 1,859 | 26.8 (25.0 to 28.5) | +1.1 |
| Activation state | Already entered or returned | Outcome and reason block not imputed | 704 | 11.3 (9.8 to 12.9) | -3.3 |
| Activation state | Already entered or returned | Reason block not imputed | 719 | 11.4 (9.9 to 13.0) | -3.2 |
| Activation state | Already entered or returned | All formal-model records | 973 | 14.6 (13.2 to 16.0) | +0.0 |
| Activation state | Already entered or returned | Outcome not imputed | 812 | 13.7 (12.0 to 15.3) | -0.9 |
| Reason prevalence | retirement | Outcome and reason block not imputed | 3,761 | 48.5 (46.5 to 50.5) | +6.1 |
| Reason prevalence | retirement | Outcome not imputed | 3,804 | 47.0 (45.1 to 48.9) | +4.7 |
| Reason prevalence | retirement | Reason block not imputed | 3,806 | 48.1 (46.2 to 50.1) | +5.8 |
| Reason prevalence | retirement | All formal-model records | 3,988 | 42.3 (40.7 to 44.0) | +0.0 |
| Reason prevalence | family_caregiving | Outcome and reason block not imputed | 1,356 | 21.8 (19.9 to 23.8) | -0.4 |
| Reason prevalence | family_caregiving | All formal-model records | 1,571 | 22.2 (20.4 to 24.0) | +0.0 |
| Reason prevalence | family_caregiving | Outcome not imputed | 1,393 | 21.5 (19.6 to 23.4) | -0.7 |
| Reason prevalence | family_caregiving | Reason block not imputed | 1,358 | 21.6 (19.6 to 23.5) | -0.6 |
| Reason prevalence | disability_or_illness | Reason block not imputed | 675 | 10.3 (9.2 to 11.4) | -0.6 |
| Reason prevalence | disability_or_illness | Outcome and reason block not imputed | 665 | 10.4 (9.2 to 11.5) | -0.5 |
| Reason prevalence | disability_or_illness | All formal-model records | 779 | 10.9 (9.8 to 12.0) | +0.0 |
| Reason prevalence | disability_or_illness | Outcome not imputed | 684 | 10.4 (9.3 to 11.5) | -0.5 |
| Reason prevalence | covid_risk_health_condition | Outcome not imputed | 382 | 5.5 (4.6 to 6.5) | -0.1 |
| Reason prevalence | covid_risk_health_condition | Reason block not imputed | 368 | 5.4 (4.5 to 6.3) | -0.3 |
| Reason prevalence | covid_risk_health_condition | Outcome and reason block not imputed | 368 | 5.5 (4.5 to 6.5) | -0.2 |
| Reason prevalence | covid_risk_health_condition | All formal-model records | 418 | 5.7 (4.7 to 6.6) | +0.0 |
| Reason prevalence | physical_demands | Outcome and reason block not imputed | 601 | 11.0 (9.9 to 12.2) | -0.5 |
| Reason prevalence | physical_demands | Outcome not imputed | 626 | 11.1 (10.1 to 12.2) | -0.4 |
| Reason prevalence | physical_demands | Reason block not imputed | 605 | 10.9 (9.8 to 12.0) | -0.7 |
| Reason prevalence | physical_demands | All formal-model records | 721 | 11.5 (10.4 to 12.6) | +0.0 |
| Reason prevalence | burnout | Reason block not imputed | 1,231 | 18.9 (17.5 to 20.2) | -1.4 |
| Reason prevalence | burnout | Outcome and reason block not imputed | 1,220 | 19.0 (17.6 to 20.4) | -1.3 |
| Reason prevalence | burnout | Outcome not imputed | 1,272 | 19.3 (18.0 to 20.6) | -1.0 |
| Reason prevalence | burnout | All formal-model records | 1,475 | 20.3 (18.9 to 21.7) | +0.0 |
| Reason prevalence | stressful_environment | Outcome not imputed | 1,127 | 17.4 (15.7 to 19.1) | -0.7 |
| Reason prevalence | stressful_environment | All formal-model records | 1,287 | 18.2 (16.4 to 20.0) | +0.0 |
| Reason prevalence | stressful_environment | Outcome and reason block not imputed | 1,087 | 17.4 (15.6 to 19.2) | -0.7 |
| Reason prevalence | stressful_environment | Reason block not imputed | 1,092 | 17.2 (15.4 to 18.9) | -1.0 |
| Reason prevalence | inadequate_staffing | Outcome not imputed | 783 | 15.2 (13.8 to 16.6) | -0.5 |
| Reason prevalence | inadequate_staffing | Outcome and reason block not imputed | 748 | 15.1 (13.7 to 16.5) | -0.6 |
| Reason prevalence | inadequate_staffing | Reason block not imputed | 756 | 14.9 (13.5 to 16.3) | -0.8 |
| Reason prevalence | inadequate_staffing | All formal-model records | 924 | 15.7 (14.3 to 17.1) | +0.0 |
| Reason prevalence | scheduling | All formal-model records | 755 | 11.2 (9.8 to 12.5) | +0.0 |
| Reason prevalence | scheduling | Outcome not imputed | 658 | 11.0 (9.6 to 12.4) | -0.1 |
| Reason prevalence | scheduling | Reason block not imputed | 631 | 10.7 (9.3 to 12.2) | -0.4 |
| Reason prevalence | scheduling | Outcome and reason block not imputed | 630 | 10.9 (9.4 to 12.3) | -0.3 |
| Reason prevalence | low_pay | All formal-model records | 614 | 9.6 (8.1 to 11.2) | +0.0 |
| Reason prevalence | low_pay | Outcome not imputed | 512 | 8.9 (7.3 to 10.6) | -0.7 |
| Reason prevalence | low_pay | Reason block not imputed | 486 | 8.8 (7.1 to 10.5) | -0.9 |
| Reason prevalence | low_pay | Outcome and reason block not imputed | 484 | 8.9 (7.2 to 10.6) | -0.8 |
| Reason prevalence | poor_management | Outcome and reason block not imputed | 777 | 12.2 (11.0 to 13.5) | -1.0 |
| Reason prevalence | poor_management | Outcome not imputed | 812 | 12.3 (11.1 to 13.5) | -0.9 |
| Reason prevalence | poor_management | All formal-model records | 943 | 13.2 (12.0 to 14.4) | +0.0 |
| Reason prevalence | poor_management | Reason block not imputed | 780 | 12.0 (10.8 to 13.3) | -1.2 |
| Reason prevalence | poor_collaboration | Outcome not imputed | 275 | 4.2 (3.3 to 5.1) | -0.3 |
| Reason prevalence | poor_collaboration | All formal-model records | 321 | 4.6 (3.6 to 5.5) | +0.0 |
| Reason prevalence | poor_collaboration | Outcome and reason block not imputed | 264 | 4.2 (3.3 to 5.2) | -0.3 |
| Reason prevalence | poor_collaboration | Reason block not imputed | 265 | 4.2 (3.3 to 5.1) | -0.4 |
| Reason prevalence | limited_advancement | Outcome and reason block not imputed | 176 | 3.1 (2.1 to 4.0) | -0.0 |
| Reason prevalence | limited_advancement | Reason block not imputed | 177 | 3.0 (2.1 to 4.0) | -0.1 |
| Reason prevalence | limited_advancement | All formal-model records | 216 | 3.1 (2.2 to 4.0) | +0.0 |
| Reason prevalence | limited_advancement | Outcome not imputed | 184 | 3.1 (2.2 to 4.0) | -0.1 |
| Reason prevalence | high_risk_conditions | Outcome and reason block not imputed | 572 | 10.1 (8.8 to 11.4) | -0.5 |
| Reason prevalence | high_risk_conditions | Outcome not imputed | 593 | 10.1 (8.9 to 11.4) | -0.4 |
| Reason prevalence | high_risk_conditions | Reason block not imputed | 577 | 9.9 (8.7 to 11.2) | -0.6 |
| Reason prevalence | high_risk_conditions | All formal-model records | 686 | 10.6 (9.3 to 11.8) | +0.0 |
| Reason prevalence | unsatisfactory_safety_protocol | Outcome and reason block not imputed | 305 | 5.8 (4.6 to 7.1) | -0.3 |
| Reason prevalence | unsatisfactory_safety_protocol | All formal-model records | 385 | 6.1 (5.0 to 7.3) | +0.0 |
| Reason prevalence | unsatisfactory_safety_protocol | Outcome not imputed | 322 | 5.9 (4.7 to 7.1) | -0.2 |
| Reason prevalence | unsatisfactory_safety_protocol | Reason block not imputed | 308 | 5.8 (4.5 to 7.0) | -0.4 |
| Reason prevalence | workplace_harassment_or_violence | Reason block not imputed | 225 | 3.9 (3.0 to 4.8) | -0.1 |
| Reason prevalence | workplace_harassment_or_violence | Outcome and reason block not imputed | 224 | 3.9 (3.0 to 4.8) | +0.0 |
| Reason prevalence | workplace_harassment_or_violence | All formal-model records | 272 | 3.9 (3.0 to 4.8) | +0.0 |
| Reason prevalence | workplace_harassment_or_violence | Outcome not imputed | 238 | 3.9 (3.0 to 4.8) | +0.0 |
| Reason prevalence | liability_concern | Outcome not imputed | 346 | 5.9 (4.8 to 6.9) | -0.3 |
| Reason prevalence | liability_concern | Reason block not imputed | 333 | 5.9 (4.8 to 6.9) | -0.3 |
| Reason prevalence | liability_concern | Outcome and reason block not imputed | 331 | 5.9 (4.9 to 7.0) | -0.3 |
| Reason prevalence | liability_concern | All formal-model records | 402 | 6.2 (5.2 to 7.2) | +0.0 |
| Reason prevalence | inability_to_practice_professionally | Outcome and reason block not imputed | 136 | 1.9 (1.3 to 2.4) | -0.3 |
| Reason prevalence | inability_to_practice_professionally | Outcome not imputed | 144 | 2.1 (1.4 to 2.8) | -0.1 |
| Reason prevalence | inability_to_practice_professionally | Reason block not imputed | 136 | 1.8 (1.3 to 2.4) | -0.4 |
| Reason prevalence | inability_to_practice_professionally | All formal-model records | 160 | 2.2 (1.6 to 2.8) | +0.0 |
| Reason prevalence | career_change | Reason block not imputed | 623 | 11.8 (10.2 to 13.3) | -0.7 |
| Reason prevalence | career_change | Outcome and reason block not imputed | 616 | 11.8 (10.3 to 13.4) | -0.7 |
| Reason prevalence | career_change | Outcome not imputed | 628 | 11.6 (10.1 to 13.2) | -0.9 |
| Reason prevalence | career_change | All formal-model records | 763 | 12.5 (11.1 to 14.0) | +0.0 |
| Reason prevalence | school_or_education | Outcome not imputed | 156 | 3.3 (2.5 to 4.2) | -0.8 |
| Reason prevalence | school_or_education | All formal-model records | 211 | 4.2 (3.2 to 5.2) | +0.0 |
| Reason prevalence | school_or_education | Outcome and reason block not imputed | 138 | 2.9 (2.2 to 3.7) | -1.2 |
| Reason prevalence | school_or_education | Reason block not imputed | 141 | 2.9 (2.2 to 3.7) | -1.3 |
| Reason prevalence | outdated_skills | Outcome not imputed | 247 | 3.7 (3.1 to 4.2) | +0.1 |
| Reason prevalence | outdated_skills | Outcome and reason block not imputed | 234 | 3.6 (3.0 to 4.1) | +0.0 |
| Reason prevalence | outdated_skills | Reason block not imputed | 236 | 3.5 (3.0 to 4.0) | -0.0 |
| Reason prevalence | outdated_skills | All formal-model records | 275 | 3.6 (3.0 to 4.1) | +0.0 |
| Reason prevalence | difficulty_finding_position | All formal-model records | 246 | 3.1 (2.1 to 4.0) | +0.0 |
| Reason prevalence | difficulty_finding_position | Outcome not imputed | 223 | 2.6 (1.8 to 3.4) | -0.5 |
| Reason prevalence | difficulty_finding_position | Reason block not imputed | 216 | 2.6 (1.7 to 3.4) | -0.5 |
| Reason prevalence | difficulty_finding_position | Outcome and reason block not imputed | 214 | 2.6 (1.8 to 3.4) | -0.5 |
| Reason prevalence | other | All formal-model records | 419 | 6.1 (5.0 to 7.2) | +0.0 |
| Reason prevalence | other | Outcome not imputed | 340 | 5.4 (4.2 to 6.5) | -0.7 |
| Reason prevalence | other | Reason block not imputed | 347 | 5.5 (4.4 to 6.7) | -0.6 |
| Reason prevalence | other | Outcome and reason block not imputed | 316 | 4.8 (3.8 to 5.9) | -1.3 |

**Supplementary Table S5 | Survey-weighted phi correlations among the ten modeled reasons**

Values were calculated in the 7,744 model-eligible respondents using the NSSRN final person weight.

| **Reason** | **Retirement** | **Family** | **Health** | **Burnout** | **Stress** | **Staffing** | **Scheduling** | **Pay** | **Management** | **Safety** |
| --- | --- | --- | --- | --- | --- | --- | --- | --- | --- | --- |
| Retirement | 1.00 | -0.29 | -0.16 | -0.22 | -0.19 | -0.17 | -0.16 | -0.20 | -0.15 | -0.18 |
| Family | -0.29 | 1.00 | -0.10 | 0.02 | 0.03 | 0.03 | 0.09 | -0.02 | -0.02 | 0.03 |
| Health | -0.16 | -0.10 | 1.00 | 0.21 | 0.31 | 0.26 | 0.22 | 0.09 | 0.20 | 0.23 |
| Burnout | -0.22 | 0.02 | 0.21 | 1.00 | 0.58 | 0.53 | 0.34 | 0.31 | 0.46 | 0.39 |
| Stress | -0.19 | 0.03 | 0.31 | 0.58 | 1.00 | 0.57 | 0.42 | 0.33 | 0.55 | 0.50 |
| Staffing | -0.17 | 0.03 | 0.26 | 0.53 | 0.57 | 1.00 | 0.43 | 0.36 | 0.59 | 0.52 |
| Scheduling | -0.16 | 0.09 | 0.22 | 0.34 | 0.42 | 0.43 | 1.00 | 0.30 | 0.35 | 0.28 |
| Pay | -0.20 | -0.02 | 0.09 | 0.31 | 0.33 | 0.36 | 0.30 | 1.00 | 0.32 | 0.27 |
| Management | -0.15 | -0.02 | 0.20 | 0.46 | 0.55 | 0.59 | 0.35 | 0.32 | 1.00 | 0.49 |
| Safety | -0.18 | 0.03 | 0.23 | 0.39 | 0.50 | 0.52 | 0.28 | 0.27 | 0.49 | 1.00 |

**Supplementary Table S6 | Machine-readable result files**

| **File** | **Contents** |
| --- | --- |
| Table_S1_characteristics_numeric.csv | Long-format numeric values underlying Table 1 |
| Table_S2_reason_state_cells.csv | Complete reason-by-activation-state cell counts and weighted estimates |
| Table_S3_multinomial_RRR.csv | Relative risk ratios for the primary and all sensitivity models |
| Table_S4_sensitivity_joint_tests.csv | Finite-design and asymptotic reason-level joint tests |
| Table_S5_imputation_comparison.csv | Full imputation comparison for activation states and reasons |
| Table_S6_weighted_reason_correlations.csv | Weighted phi correlation matrix |
